# MRI assessment of adipose tissue fatty acid composition in the UK Biobank and its association with diet and disease

**DOI:** 10.1101/2024.03.27.24304957

**Authors:** Marjola Thanaj, Nicolas Basty, Brandon Whitcher, Jimmy D Bell, E Louise Thomas

## Abstract

**Objectives:** This study aimed to assess the fatty acid (FA) composition of abdominal subcutaneous (ASAT) and visceral (VAT) adipose tissue in the UK Biobank imaging cohort (N = 33,823) using magnetic resonance imaging (MRI).

**Methods:** We measured the fractions of saturated (fSFA), monounsaturated (fMUFA), and polyunsaturated (fPUFA) in ASAT and VAT from multi-echo MRI scans. We selected a sub-cohort that followed a vegan and an omnivore diet (N=36) to validate the effect of diet on adipose tissue. In the wider imaging cohort, we examined the relationship between adipose tissue FA composition and various traits related to disease and body size.

**Results:** We measured adipose tissue FA composition for over 33,000 participants, revealing higher fSFA and fPUFA and lower fMUFA in VAT (p < 0.00016). fMUFA and fPUFA were higher in ASAT and lower in VAT for women (p<0.00016). Vegans exhibited lower fSFA in both ASAT and VAT (p < 0.00016). VAT fSFA and fMUFA showed significant associations with disease as well as anthropometric variables.

**Discussion:** This extensive analysis revealed the relationships between adipose tissue FA composition and a range of factors in a diverse population, highlighting the importance of studying body adipose tissue beyond its quantity.

**Study importance:** *What is already known?:*

- The fatty acid (FA) composition of adipose tissue is an independent risk factor for hypertension, type-2 diabetes (T2D) and cardiovascular disease.
- There has yet to be a large-scale population study of adipose tissue FA composition, principally due to the invasive nature of available methods.

*What does the study add?:*

- We show that MRI-based methods can be readily applied across a large population (n=33,823) while confirming and expanding on the associations between dietary patterns and FA composition in both abdominal subcutaneous (ASAT) and visceral (VAT) adipose tissue.
- Models involving saturated and monounsaturated FA composition in VAT demonstrate significant association with disease outcomes, anthropometric variables, dietary macronutrient intake and physical activity.

*How might these results change the direction of research or the focus of clinical practice?:*

- Our findings highlight the importance of evaluating adipose tissue composition and its relationship with dietary and disease traits.
- These insights will contribute to formulating more informed lifestyle recommendations to alleviate or even reverse metabolic conditions associated with obesity.

## Introduction

While adipose tissue content and distribution are strongly associated with the development of metabolic disease, adipose tissue composition, a reflection of long-term dietary fatty acid (FA) intake, is also thought to play a significant role (1). Indeed, the relative proportions of dietary saturated (SFA), unsaturated (UFA), monounsaturated (MUFA) and polyunsaturated (PUFA) FA are associated with the development of type 2 diabetes (2), hypertension (3), and cardiovascular disease (CVD) (4).

Adipose tissue FA composition reflects long-term dietary and habitual FA intake, with gas-liquid chromatography (GLC) of adipose tissue biopsies being the gold standard for its measurement (5). However, the invasivity of this method limits the sample size and the anatomical location that can be studied. Non-invasive methods such as *in vivo* carbon-13 (^13^C) and proton (^1^H) magnetic resonance spectroscopy (MRS) have been used to measure human adipose tissue FA composition (6). Although these methods cannot measure individual fatty acids like GLC, they estimate the relative proportions of SFAs, UFAs, MUFAs and PUFAs. Thus, they have been applied to the study of adipose tissue FA composition in relation to diet (7), development (8) and disease (9). However, these non-invasive methods have not been widely adopted despite their potential, mainly due to the considerable technical demands, significantly limiting their applicability. Recently, Hamilton et al. (10) developed a ^1^H MRS method to characterise the liver fat FA composition, later applied by Bydder et al. (11) to estimate adipose tissue composition from magnetic resonance imaging (MRI). This approach of measuring adipose tissue FA composition is increasingly being used (11–13), partly due to its applicability on most MRI scanners, providing estimates for the number of double bonds (NDB), the number of methylene-interrupted double bonds (NMIDB) and the FA chain length (10). These parameters enable the derivation of fractions of saturated (fSFA), unsaturated (fUFA), monounsaturated (fMUFA) and polyunsaturated (fPUFA) fatty acids, which have been validated against GLC and MRS (14).

Besides being widely available, MRI assessment of adipose tissue composition provides enhanced spatial information, allowing simultaneous estimation across multiple adipose tissue depots and sites such as the liver, muscle and bone marrow (15). This enables the detection of differences in FA composition between VAT and ASAT depots and between superficial and deep layers of ASAT (16).

Previous studies have demonstrated that FA composition of adipose tissue influences the risk of various diseases, such as insulin resistance (17), osteoporosis (18), and all-cause mortality (19). However, MRI assessment of adipose tissue composition in large cohorts has been scarce, with most studies involving only a few tens or hundreds of participants, reflecting challenges in scaling up these methods (12).

Our study aimed to test the feasibility of using MRI to measure adipose tissue composition in a large cohort of participants and determine the FA composition of VAT and ASAT. We also sought to confirm previous observations regarding the relationship between ASAT and VAT FA composition and habitual diet. We finally explored the effect of adipose tissue FA composition on factors such as lifestyle and disease, including hypertension, T2D and CVD.

## Methods

### Data

Images were obtained using a Siemens Aera 1.5T scanner (Syngo MR D13) (Siemens, Erlangen, Germany), with full details of the UK Biobank abdominal MRI protocol previously reported (20). Our analysis used the neck-to-the-knee chemical-shift-based images, and the pancreas single-slice multi-echo sequence in 33,823 participants. SAT and VAT segmentations were obtained from the three-dimensional chemical-shift-encoded MRI sequences using a deep learning algorithm (21) and projected into the single-slice multi-echo data.

Participant data from the UK Biobank cohort was obtained through UK Biobank Access Application number 44584. The UK Biobank has approval from the North West Multi-Centre Research Ethics Committee (REC reference: 11/NW/0382), with informed consent obtained from all participants. Researchers may apply to use the UKBB data resource by submitting a health-related research proposal in the public interest. Additional information may be found on the UK Biobank researchers and resource catalogue pages (http://www.ukbiobank.ac.uk).

### Phenotype Definitions

We used anthropometric measurements, including age, body mass index (BMI), waist and hip circumference, obtained at the UK Biobank imaging assessment. We also used self-reported ethnicity and sex, as well as Townsend deprivation index from the initial UK Biobank assessment visit. We calculated participant’s excess metabolic equivalents of task (MET) in hours per week by multiplying the time in minutes spent in each physical activity, including walking, moderate and vigorous activity, by the number of reported days doing the exercise and the respective MET scores using previously described methods (22). We used the vigorous MET as a more reliable measure of intensive physical activity. All physical activity measures, alcohol intake frequency and smoking, were self-reported at the UK Biobank imaging visit. Inflammatory biomarkers, including c-reactive protein (CRP) and white blood cell count (WBCC) were obtained at the initial assessment visit.

UK Biobank dietary information was collected using the Oxford WebQ (www.ceu.ox.ac.uk/research/oxford-webq), a web-based recall questionnaire for large population studies. We calculated mean values from the data of the five dietary assessments on which UK Biobank participants completed the questionnaire. We estimated the dietary intake of PUFAs by subtracting SFAs and MUFAs intake from total fat intake. The primary exposure variables were the dietary intake derived from the macronutrients, including carbohydrates, fat, and protein, and components, including MUFA, PUFA, and SFA in grams as well as the energy intake (in kJ). In our following analysis, all dietary intake from the FA components will be referred to as dietary MUFA, PUFA, and SFA.

Given that previous studies have shown, using *in vivo* ^13^C MRS, that vegan participants show significant differences in adipose tissue composition, compared to omnivores (23), we studied a subcohort of vegan participants as proof of principle for our methods. Vegan participants were defined by self-reported, at the first imaging visit, habitual “vegan diet” or “never” consuming meat, fish, dairy, and eggs, whereas omnivores were selected by self-reporting “once a week” consuming meat, fish, dairy, and egg intake (24). Relevant conditions of interest, including hypertension, T2D and CVD, were defined at the time and before the imaging visit. A summary of the codes corresponding to the diet and disease condition considered is shown in Supplementary Table S1.

### Data Analysis

The MRI-based method used to quantify adipose tissue FA composition in this study is based on Bydder et al. (11), where the NDB, NMIDB, and chain length describe triglyceride molecules. In brief, NDB were determined iteratively by fitting the MR signal in a nonlinear least-square approach as a function of the echo times. The NMIDB were calculated from the NDB using a previously described heuristic technique (NMIDB = 0.093 × NDB²) (11). Bound constraints for NDB were 1 ≤ NDB ≤ 6.

FA Composition was derived from NDB and NMIDB using the following relations (25):

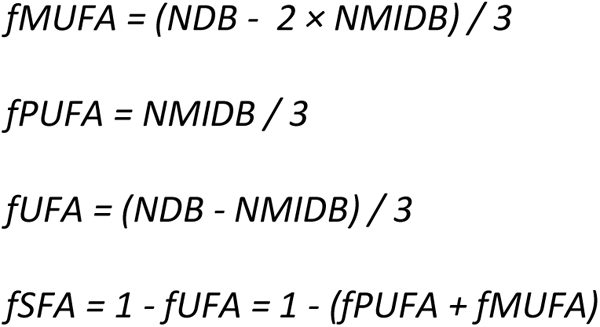

Where fUFA is the UFA fraction, fSFA is the SFA fraction, fMUFA is the MUFA fraction, and fPUFA is the PUFA fraction. All the estimated adipose tissue FA composition values fell within the range of 0 to 1. Estimates of NDB, NMIDB, fUFA, fMUFA, fPUFA and fSFA were calculated voxel-by-voxel, creating FA composition maps.

Segmentations of SAT and VAT were used to isolate these two adipose tissue compartments on the FA composition maps. Post-processing included one iteration of binary erosion to the ASAT and VAT masks to remove voxels that suffer from partial-volume effects where non-adipose tissue may be present. Voxels within these masks with an estimated fat fraction below 20% were also removed, as suggested by Bydder et al. (11). Our analysis will mainly focus on fMUFA, fPUFA and fSFA values to describe FA composition in adipose tissue.

### Quality Control

After post-processing of ASAT and VAT segmentations, 197 participants were excluded since no voxels remained in the fat depot masks, which was confirmed by visual inspection. We further conducted quality control measures to determine potential outliers in the estimated NDB values by visually examining the lowest and highest values of their respective ranges (i.e. 1 > NDB > 6), resulting in the exclusion of an additional 43 participants. For consistency in the sample size, participants with missing data for one fat depot (e.g. ASAT) were excluded from the study even if the other fat depot (e.g. VAT) had full coverage. From the initial 33,823 participants, 33,583 were included in our analysis (0.7% of the data excluded). A flow diagram of the inclusion/exclusions in the study population is shown in Supplementary Figure S1.

### Statistical Analysis

Descriptive characteristics are presented as means and standard deviations (SDs) for quantitative variables and as counts for categorical variables using the R software environment (26). The Wilcoxon rank-sum test was used to compare means between groups, and the Wilcoxon signed-rank test was used for groups with paired observations. We assessed the effect of adipose tissue FA composition on vegans and omnivores after adjusting for age, sex, alcohol, smoking, MET and the Townsend deprivation index, using linear regression models using the adipose tissue FA composition measurements as dependent variables in each analysis. BMI was excluded from the linear regression analysis due to its high correlation with FA composition in both ASAT and VAT. In the full imaging cohort, we evaluated the impact of adipose tissue FA composition on anthropometric, lifestyle and disease factors using linear regression models, adjusting for age, sex, ethnicity, BMI, alcohol, smoking, vigorous MET, the Townsend deprivation index, intake from macronutrients including fat, carbohydrate and protein and their components including dietary MUFA, PUFA and SFA, hypertension, T2D and CVD. To avoid issues related to collinearity among dietary variables, we excluded the intake of dietary PUFA and dietary fat from our model. In our linear regression analyses, we excluded all participants who, at any of the five dietary assessments, reported that what they ate or drank yesterday was not typical (27). All continuous variables were standardised, with units in one standard deviation (SD), before being included in the linear regression models. Summaries of the linear regression analysis are reported as standardised regression coefficients (β) with standard errors in the tables and as standardised regression coefficients with 95% confidence intervals in the figures. After counting the hypothesis tests, the Bonferroni-corrected threshold for statistical significance was 0.05/309 = 0.00016.

## Results

### Baseline and Adipose Tissue Composition Characteristics

Baseline characteristics and summary statistics of the whole cohort, including adipose tissue FA composition, are provided in Tables 1, and 2, and separated by sex in Supplementary Table S2. Of the 33,583 participants, 96.8% were of White ethnicity, and 51.4% were women, with an average age of 63.80 ± 7.50 years for women and 65.17 ± 7.80 years for men. There was a very small, yet significant, sex difference in BMI (women: 26.04 ± 4.64 kg/m² vs men 26.89 ± 3.74 kg/m², p < 0.00016). Reported vigorous MET values were significantly lower for women than men participants (6.56 ± 7.75 hours/week vs 7.84 ± 8.20 hours/week). Men had higher dietary MUFA, PUFA and SFA, carbohydrates and protein intake, compared to women (p < 0.00016). Within the study cohort, 12,093 participants had hypertension, 1,765 were diagnosed with T2D, and 3,306 were diagnosed with CVD. fSFA and fPUFA were higher in VAT, whereas fMUFA were lower in VAT than in ASAT (p < 0.00016) for both men and women. ASAT fMUFA and fPUFA were significantly higher, and fSFA was significantly lower in women than in men. Conversely, VAT fMUFA and fPUFA were significantly lower, and fSFA was significantly higher in women than men (Table 2).

**Table 1.**
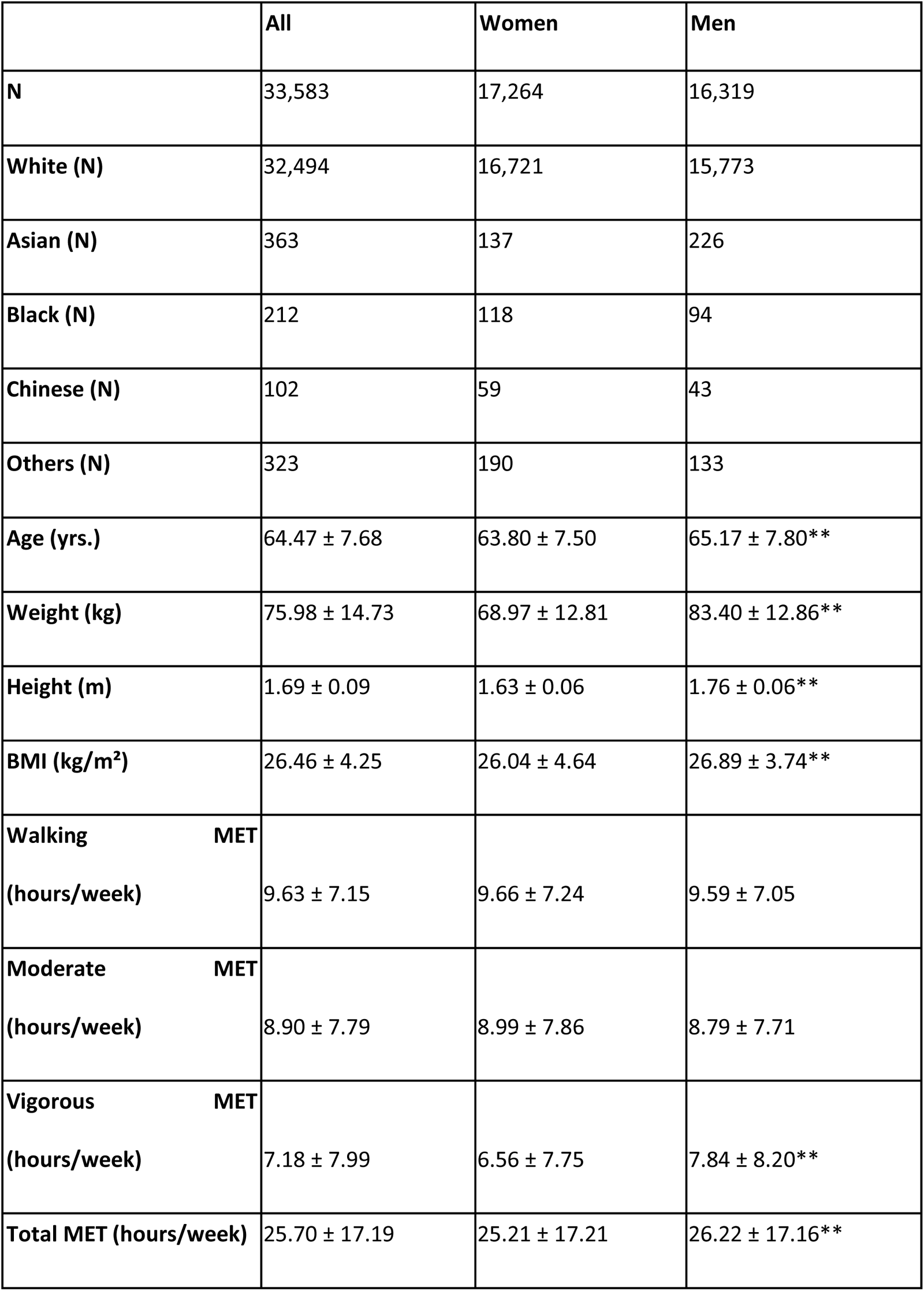
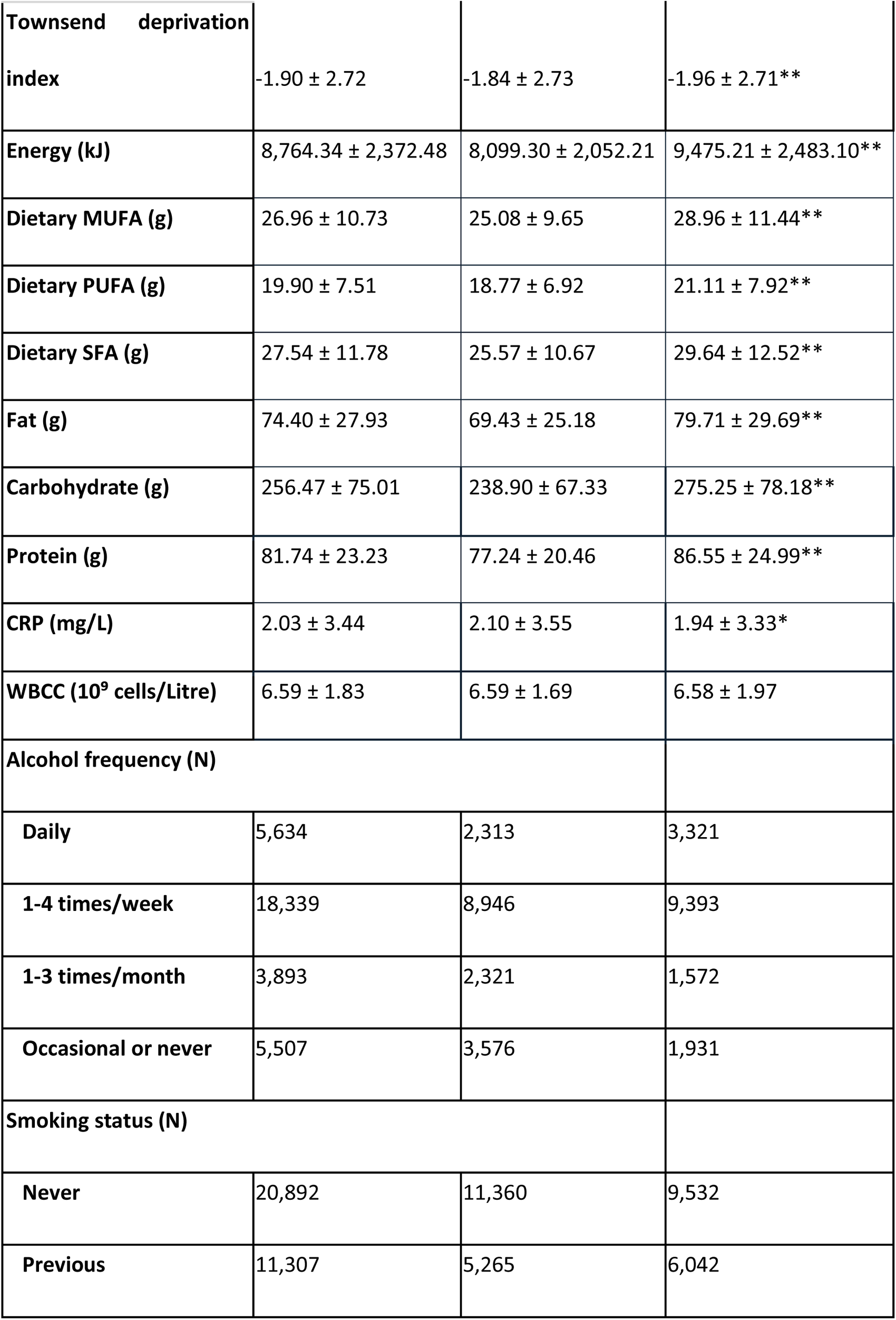
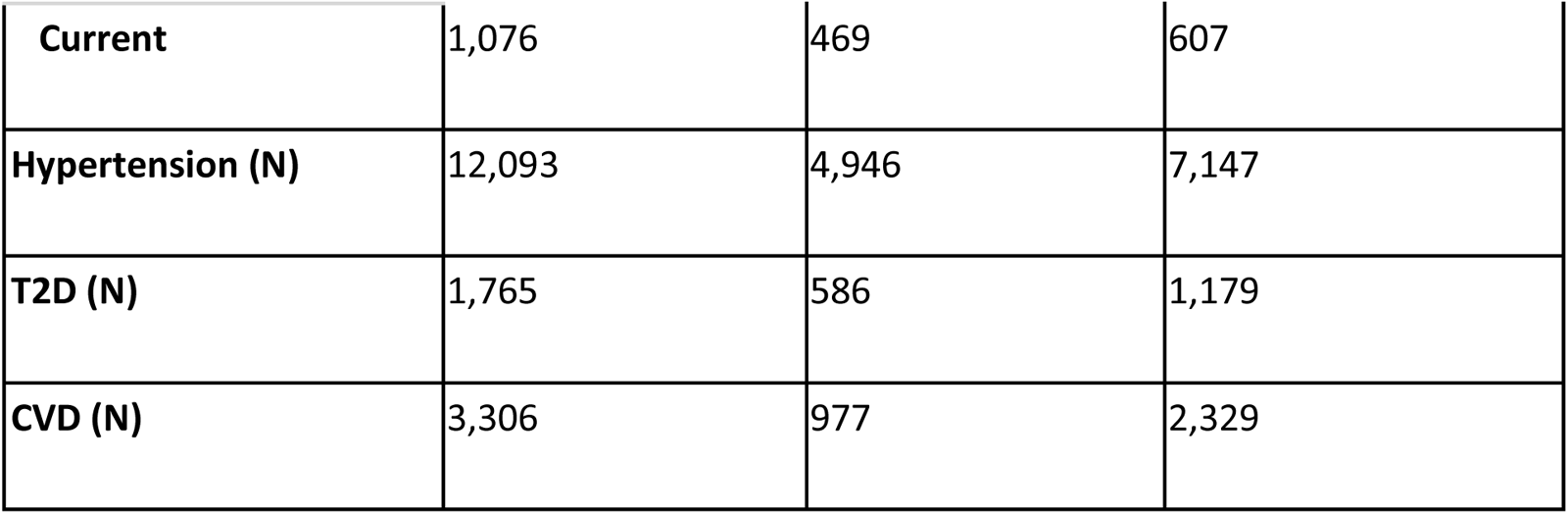
Demographics of the full cohort, separated by sex. Values are reported as mean ± standard deviation for continuous variables and counts (N) for categorical variables. Significance refers to the p-value for a Wilcoxon rank-sums test, where the null hypothesis is the medians between the two groups (women and men) are equal. An asterisk (*) indicates statistical significance for p-value < 0.05, ** indicates statistical significance after Bonferroni correction (p = 0.00016). Abbreviations: BMI, body mass index; MET, metabolic equivalents of task; FAs, fatty acids; CRP, c-reactive protein; WBCC, white blood cell count; T2D, type-2 diabetes; CVD, cardiovascular disease.

**Table 2.**
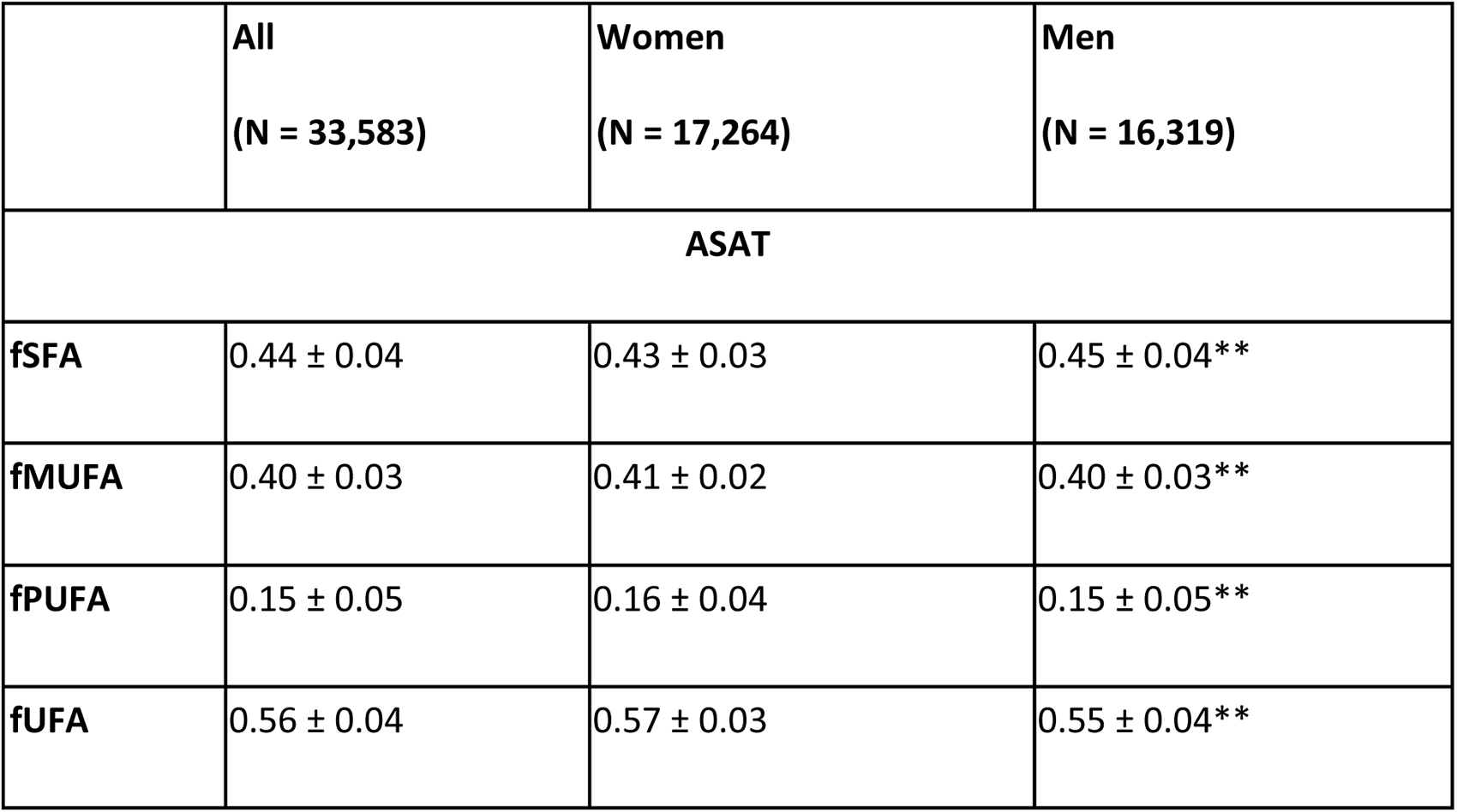
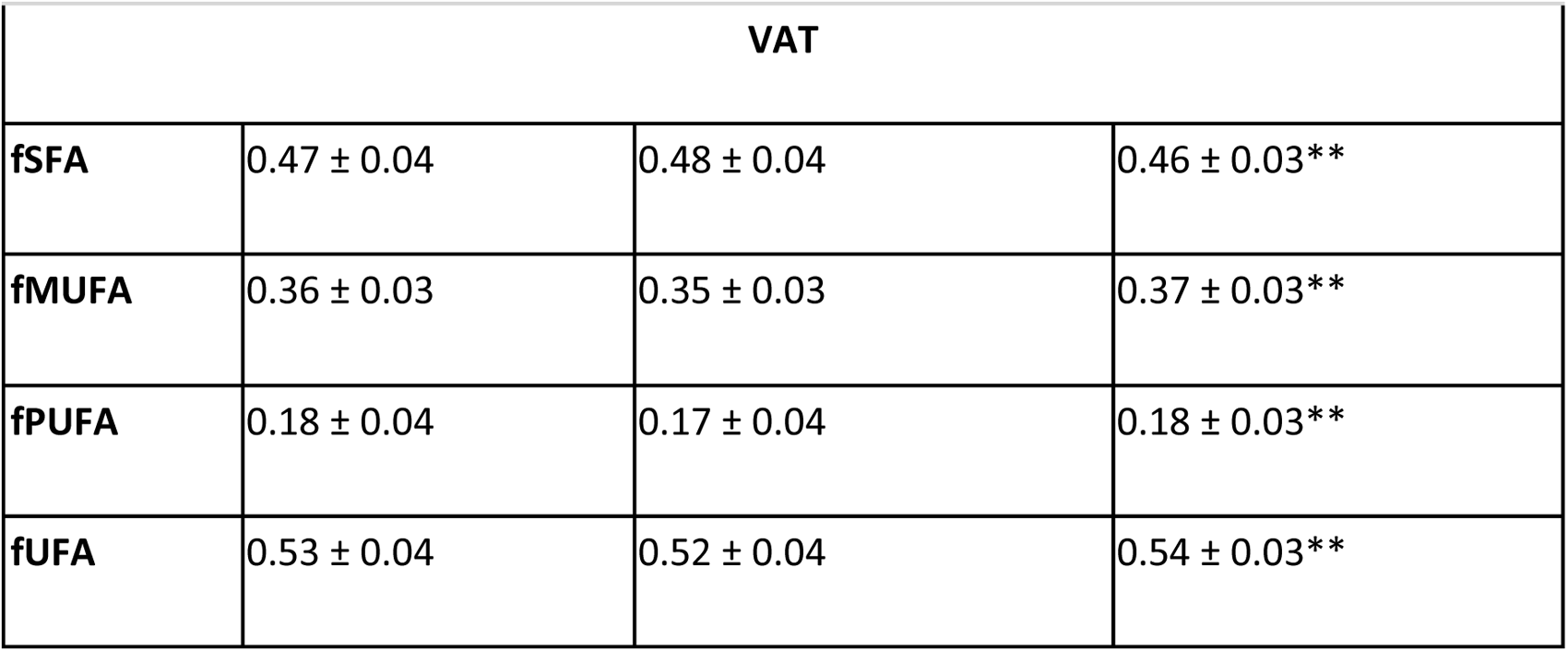
Summary statistics for the full cohort’s FA composition in ASAT and VAT, separated by sex. Values are reported as mean ± standard deviation. Significance refers to the p-value for a Wilcoxon rank-sums test, where the null hypothesis is the medians between the two groups are equal. ** indicates statistical significance between the vegan and omnivore participants, after Bonferroni correction (p = 0.00016). After Bonferroni correction (p = 0.00016), all comparisons between FA composition in ASAT and VAT, for the full cohort and separated by sex, were statistically significant. Abbreviations: fSFA, saturated fatty acid fraction; fPUFA, polyunsaturated fatty acids fraction; fMUFA, monounsaturated fatty acids fraction; fUFA, unsaturated fatty acid fraction; ASAT: abdominal subcutaneous adipose tissue; VAT: visceral adipose tissue.

To validate our adipose tissue FA composition measurements, we examined a subset of vegan and omnivore participants. This subgroup was selected as vegan participants are known to have marked differences in dietary fat intake (elevated unsaturated fat consumption), which is reflected in their adipose tissue composition. We identified 51 vegan participants in the UK Biobank, from which only 18 had complete pancreas multi-echo scans, leading us to match 18 vegans with 18 omnivores based on gender. Figure 1 illustrates an example of FA composition in ASAT and VAT for representative vegan and omnivore participants, while baseline characteristics and summary statistics of adipose tissue FA composition are provided in Tables 3 and 4, and by sex in Supplementary Tables S3 and S4.

**Figure 1.**
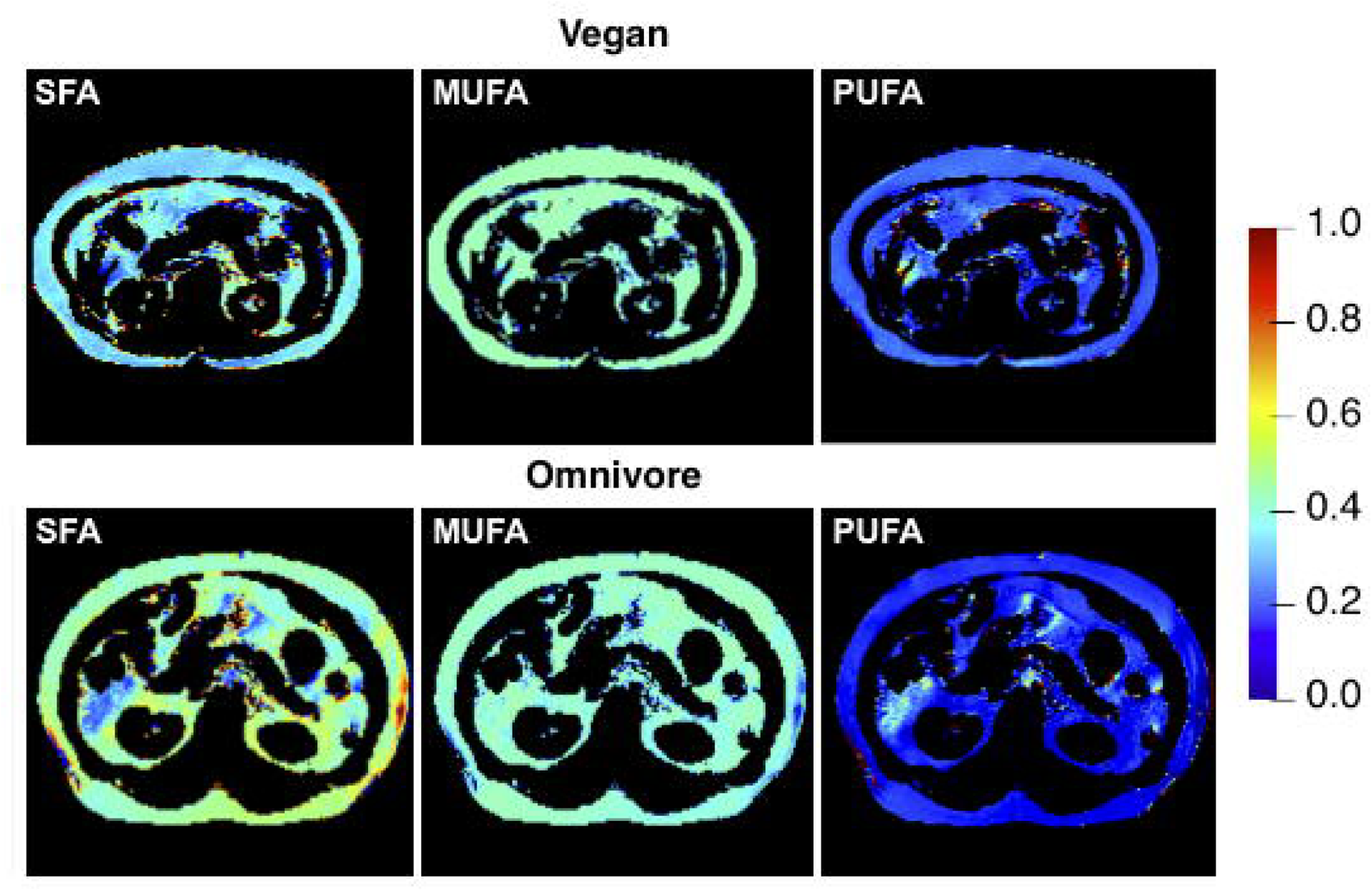
Examples of estimated fSFA, fMUFA and fPUFA maps on the ASAT and VAT of a vegan and an omnivore participant. The fSFA and fPUFA are visibly different between the two participants. Abbreviations: fSFA, saturated fatty acid fraction; fPUFA, polyunsaturated fatty acids fraction; fMUFA, monounsaturated fatty acids fraction; ASAT: Abdominal subcutaneous adipose tissue; VAT: Visceral adipose tissue.

**Table 3.** Demographics of the vegan and omnivore participants. Values are reported as mean ± standard deviation for continuous variables and counts (N) for categorical variables. Significance refers to the p-value for a Wilcoxon rank-sums test, where the null hypothesis is the medians between the two groups (vegan and omnivore participants) are equal. An asterisk (*) indicates statistical significance for p-value < 0.05, ** indicates statistical significance after Bonferroni correction (p = 0.00016). Abbreviations: BMI, body mass index; MET, metabolic equivalents of task.

|  | **Vegan** | **Omnivore** |
| --- | --- | --- |
| **N** | 18 | 18 |
| **White (N)** | 17 | 18 |
| **Age (yrs.)** | 60.94 ± 7.16 | 71.56 ± 12.20* |
| **Weight (kg)** | 67.09 ± 10.54 | 100.90 ± 18.93** |
| **Height (m)** | 1.68 ± 0.09 | 1.69 ± 0.09 |
| **BMI (kg/m<sup>2</sup>)** | 23.61 ± 2.68 | 35.22 ± 5.79** |
| **Walking MET (hours/week)** | 8.54 ± 8.44 | 8.36 ± 8.94 |
| **Moderate MET (hours/week)** | 23.26 ± 32.35 | 6.21 ± 11.74* |
| **Vigorous MET (hours/week)** | 8.13 ± 13.40 | 3.41 ± 8.58 |
| **Total MET (hours/week)** | 39.94 ± 46.48 | 17.98 ± 24.14* |
| **Townsend deprivation index** | 0.09 ± 2.79 | -2.47 ± 2.20* |
| **Alcohol frequency (N)** |  |  |
| **Daily** | 3 | 3 |
| **1-4 times per week** | 6 | 5 |
| **1-3 times per month** | 2 | 3 |
| **Occasional or never** | 7 | 7 |
| **Smoking status (N)** |  |  |
| **Never** | 13 | 11 |
| **Previous** | 4 | 6 |
| **Current** | 1 | 1 |

**Table 4.**
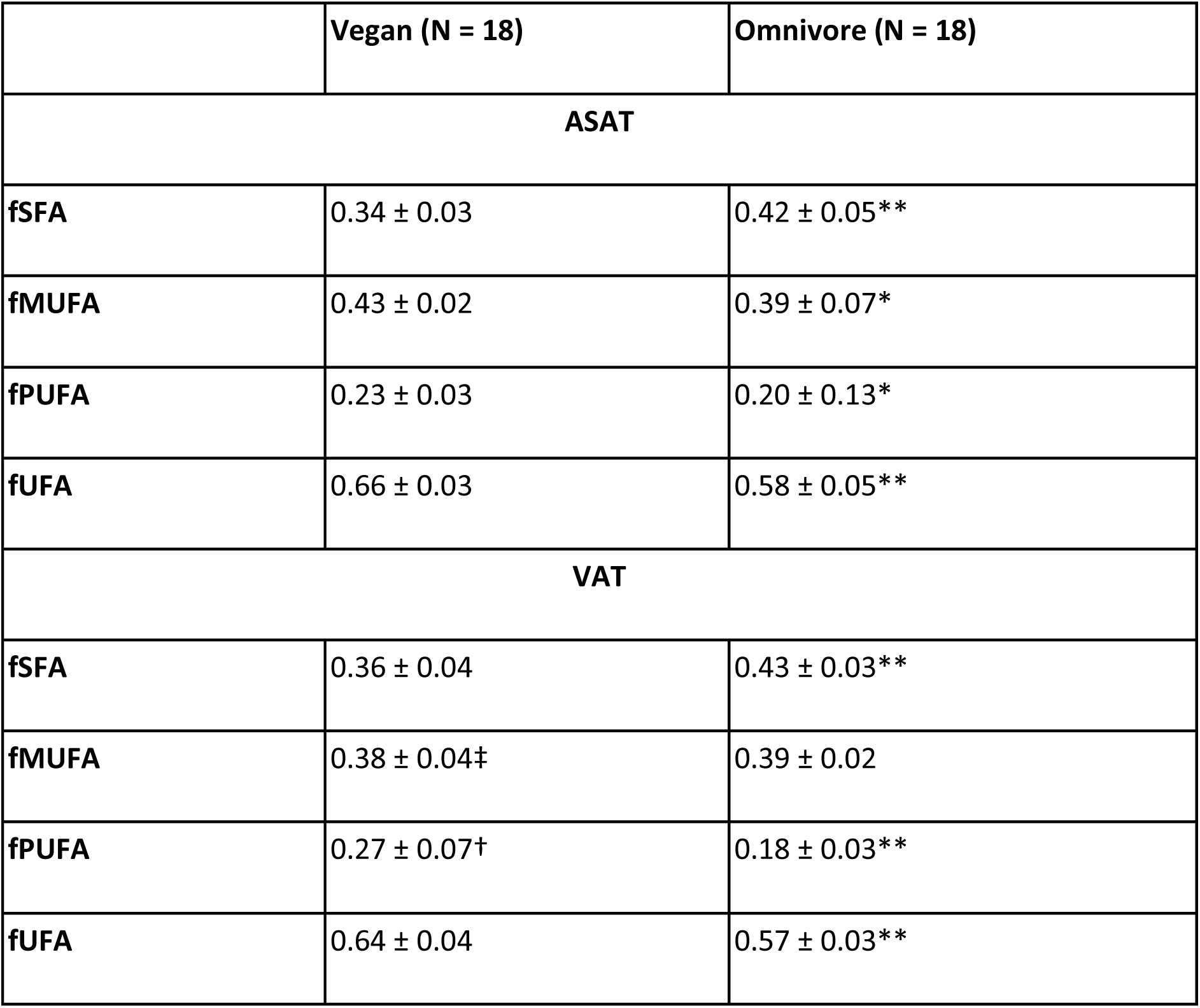
Summary statistics for FA composition in ASAT and VAT of the vegan and omnivore participants. Values are reported as mean ± standard deviation. Significance refers to the p-value for a Wilcoxon rank-sums test, where the null hypothesis is the medians between the two groups are equal. An asterisk (*) indicates statistically significant for p-value < 0.05, ** indicates statistical significance between the vegan and omnivore participants, after Bonferroni correction (p = 0.00016). † indicates statistical significance between FA composition in ASAT and VAT, for p-value < 0.05 and ‡ indicates statistical significance between FA composition in ASAT and VAT, after Bonferroni correction (p = 0.00016). Abbreviations: fSFA, saturated fatty acid fraction; fPUFA, polyunsaturated fatty acids fraction; fMUFA, monounsaturated fatty acids fraction; fUFA, unsaturated fatty acid fraction; ASAT: abdominal subcutaneous adipose tissue; VAT: visceral adipose tissue.

Vegans exhibited higher fMUFA in ASAT than VAT (p < 0.00016), with no other significant adipose tissue-specific differences in FA composition observed in vegans and omnivores. Significantly lower fSFA were observed in the ASAT of vegans compared to omnivores (0.34 ± 0.03 vs 0.42 ± 0.05, p < 0.00016, Table 2). Similarly, in VAT fSFA was significantly lower, whereas fPUFA was significantly higher in vegans compared to omnivores (p < 0.00016).

### FA Composition of Adipose Tissue, Anthropometric Characteristics Diet and Disease

A summary of the linear regression models for vegan and omnivore participants is provided in Supplementary Table S5. fSFA in VAT was positively associated with an omnivore diet with a standardised regression coefficient of 1.556 (p < 0.00016) and R² of 0.52. The estimated SD value for fSFA in VAT was 0.05, meaning, on average, consuming an omnivore diet results in a 0.078 increase in fSFA in VAT compared with a vegan diet. None of the models detected any further statistically significant associations. This could be attributed to the small number of participants available for this substudy (N=36).

Statistical linear regression models were used to investigate the effect of fSFA, fMUFA, and fPUFA in ASAT and VAT on diet, lifestyle factors and disease within the larger cohort (see Supplementary Table S6 and Figure 2). Of the initial 33,583 UK Biobank participants, 21,463 completed at least one dietary questionnaire. The exclusion of 8,132 participants whose dietary assessments indicated atypical diets and 1,203 participants with missing anthropometric, inflammatory and lifestyle data left 12,128 participants eligible for inclusion in the linear regression analysis (Supplementary Figure S1).

**Figure 2.**
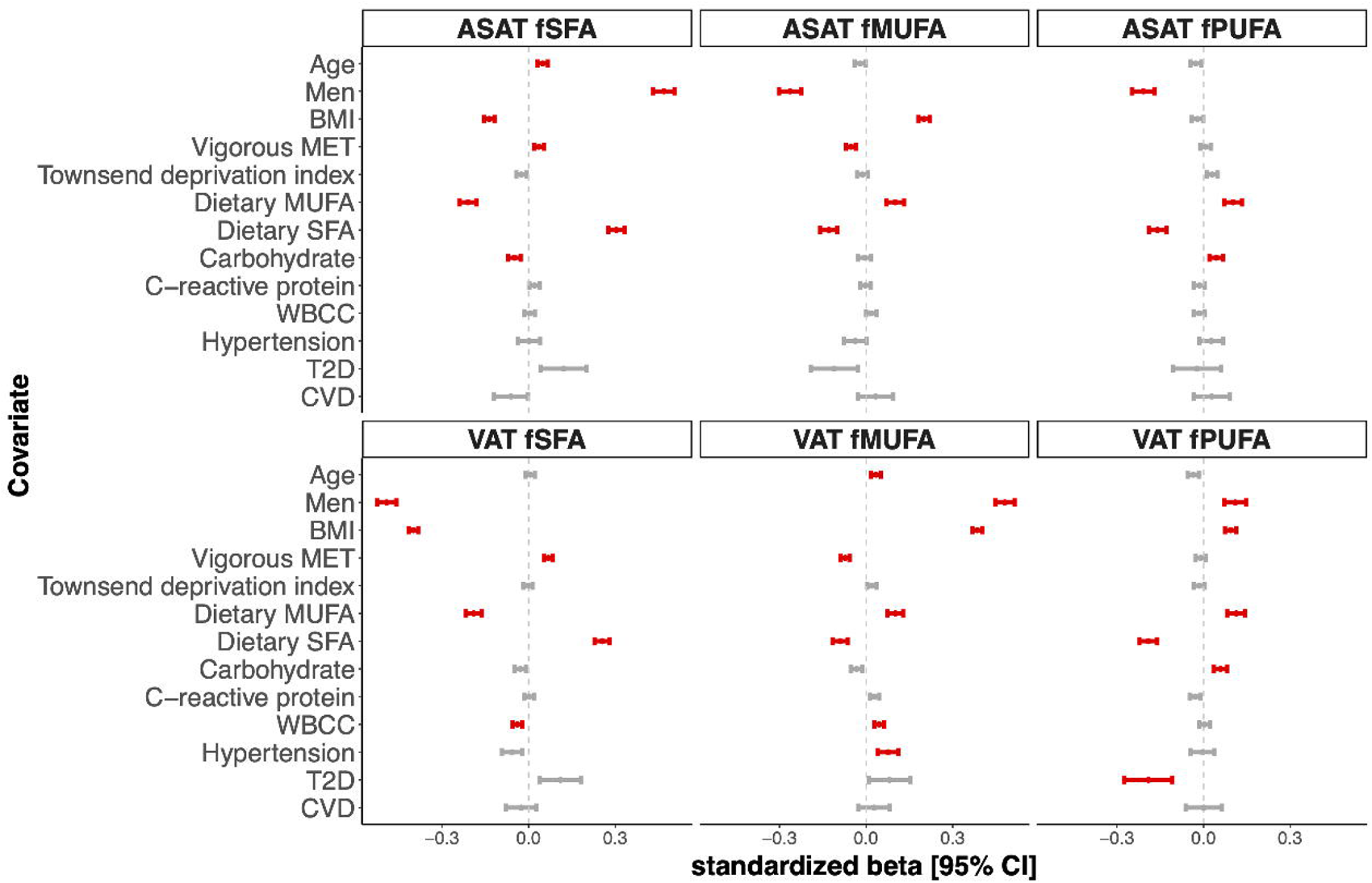
Summary of linear regression coefficients of the FA composition in ASAT and VAT representing the associations with the most relevant baseline characteristics on the full cohort. Standardised beta coefficients are displayed with 95% confidence intervals. Significant associations for p-value below the Bonferroni correction (p = 0.00016) are shown in red, and non-significant associations in grey.

fSFA in ASAT and fMUFA in VAT was positively associated with age (p < 0.00016). In ASAT, fSFA was positively associated with age and men, while fMUFA and fPUFA were negatively associated with men (fSFA: β = 0.468; fMUFA: β = -0.264; fPUFA: β = -0.209, p < 0.00016). In VAT these measures were inversely associated (p < 0.00016). BMI was negatively associated with fSFA but positively associated with fMUFA in both ASAT and VAT (p < 0.00016). fSFA in both ASAT and VAT was significantly positively associated with vigorous MET, while fMUFA was negatively associated. Dietary MUFA was negatively associated with fSFA and positively associated with fMUFA and fPUFA in both ASAT and VAT. However, dietary SFA was positively associated with fSFA and negatively associated with fMUFA, and fPUFA in ASAT and VAT (p < 0.00016). ASAT fSFA was negatively associated with carbohydrate intake, while fPUFA in both fat depots were positively associated with dietary carbohydrate intake (p < 0.00016).

In VAT, WBCC showed a significant negative association with fSFA and a positive association with fMUFA (fSFA: β = -0.039; fMUFA: β = 0.045, p < 0.00016). In the context of associations with disease, in VAT, fMUFA exhibited a positive association with hypertension (β = 0.076, p < 0.00016), while fPUFA in VAT was negatively associated with T2D (β = -0.191, p < 0.00016). It is worth noting that R^2^ values were relatively low, reflecting a poor fit in the linear regression models for all three FA compositions in both ASAT and VAT, with the highest R^2^ values being for fSFA and fMUFA in VAT (0.29 and 0.28, respectively).

## Discussion

In the present study, we estimated the FA composition of ASAT and VAT from abdominal MRI acquired from 33,823 UK Biobank participants. The increasing inclusion of abdominal MRI within national biobanks and large cohorts presents an ideal opportunity to extract maximum information from these valuable datasets. Since the UK Biobank does not collect tissue biopsies, validating our MRI findings against GLC was impossible. Therefore, we tested our overall findings by assessing our ability to detect well-established diet-related differences in adipose tissue composition, in particular those related to participants habitually following a vegan or omnivore diet. Obtaining results consistent with prior studies using ^13^C MRS, a technique validated against GLC, demonstrating higher fPUFA and lower fSFA in the adipose tissue of subjects following a vegan diet (7), gave us confidence in the reliability of our findings, enabling subsequent analyses relating to associations between diet, anthropometric, inflammatory, lifestyle and disease traits.

While numerous small human studies relating dietary fat to adipose tissue fat composition have been undertaken using GLC, very few have used MRI (14). Trinh et al. (28), reported that differences in habitual saturated fat intake are closely reflected in the FA composition of VAT and ASAT. In the present study, we confirm these findings, with dietary saturated and monounsaturated fats being positively associated with fSFA and negatively associated with fMUFA and fPUFA, respectively, in both fat depots. These results also align well with previous murine studies, where high dietary SFAs resulted in VAT containing significantly fewer MUFA and PUFA but more SFA than a diet composed primarily of PUFAs (29).

In our study, we further found that ASAT fSFA was negatively associated, while fPUFA was positively associated with dietary carbohydrates, in both ASAT and VAT. This aligns with a previous human study indicating a negative correlation between carbohydrate intake and SFA in adipose tissue composition (30). Moreover, studies in mice have suggested that high dietary carbohydrate intake is related to increased PUFA in SAT (31). A high carbohydrate diet would be expected to drive *de novo* lipogenesis, thereby increasing adipose tissue fSFA (32). However, given the relatively high dietary fat intake in the UK Biobank, it is doubtful that *de novo* lipogenesis significantly impacts adipose tissue composition. Further, research is needed to provide further insights into these dietary associations with FA composition in different fat depots and help clarify how long-term diet affects the FA profile of adipose tissue and its potential influence on metabolic outcomes.

We sought to determine the influence of sex and age on the FA composition of adipose tissue. We found that ASAT was more unsaturated in women than men, with higher levels of fMUFA and fPUFA while VAT was more saturated in women than men. In agreement with our overall findings, Machann et al. (16), using ^1^H MRS, reported ASAT to be significantly more unsaturated than VAT. However, they reported a non-significant trend for a more unsaturated adipose tissue profile in men than women. However, the same authors reported a negative association between VAT PUFA and total VAT volume (33), suggesting men with a larger VAT depot would have a more saturated VAT FA profile. Given that the differences we observed were small, the power of the large number of participants in our study may be an important factor. In our analysis of FA composition across different adipose tissues, VAT was more saturated than ASAT, whereas ASAT was more enriched in MUFA than VAT, consistently observed in both men and women, aligning with previous studies investigating FA composition differences among different human fat depots (34). However, it is worth noting that previous studies provide conflicting results, showing more MUFA and less SFA in VAT than ASAT (31). Further research may offer additional insights into these adipose tissue-specific differences.

Direct comparison of MRI results with the GLC literature is not straightforward; for instance, Lohner et al. (35), in a large meta-analysis, found certain PUFA such as linoleic acid (18:2n-6) and dihomo-γ -linolenic acid (20:3n-6) were elevated in the adipose tissue of women, while others such as gamma-linolenic acid (C18:3n-6) and arachidonic acid (C20:4n-6), and n–3 PUFAs were higher in men. There is evidence that there are sex differences in the ability to synthesise n-3 fatty acids from precursors such as alpha-linolenic acid, and sex hormones also play a role, with oestrogen stimulating and testosterone inhibiting conversion of essential FA into long-chain FAs (36). It is unclear why the sex differences between ASAT and VAT depots are inconsistent. A confounding factor may relate to the interaction between sex and age, which have both been shown to influence adipose tissue FA composition (37). In the present study, we found a significant positive association between age and adipose tissue composition only for ASAT SFA and VAT fMUFA. The variability in these patterns emphasises the sex-specific differences in ASAT and VAT regulation of the FAs pathways.

In addition to diet, sex, and ageing, multiple factors influence the FA composition of adipose tissue. We found a significant negative association between BMI and fSFA and a positive association with fMUFA in both ASAT and VAT. The reasons for these observations are unclear but may be related to various factors, including fat cell size (38), which is known to be positively associated with increasing adiposity (39).

Inflammatory biomarkers, including WBCC also showed negative associations with fSFA and positive associations with fMUFA in VAT. Previous studies suggest that SFA intake produces proinflammatory gene expression in adipose tissue, whereas MUFA intake reduces inflammatory activity (40). However, recent findings on inflammatory activity, FA intake, and red blood cell (RBC) FA in healthy humans, revealed unexpected findings, highlighting an inverse association between inflammation and total RBC SFA and a positive relationship with MUFA, attributed to an increase in a rate-limiting enzyme responsible for desaturation of both SFA and MUFA (41).

Vigorous physical activity was positively associated with fSFA and negatively associated with fMUFA in both VAT and ASAT, highlighting the potential role of exercise in modulating FA profiles within adipose tissue. Previous studies have shown that physical activity contributes to increased stearic acid (18:0) and decreased palmitoleic acid in the adipose tissue, highlighting the role of exercise in modifying adipose tissue lipid profiles and their associations with metabolic health and related diseases (42).

We found that VAT fMUFA was positively associated with hypertension. Elevated serum palmitoleic acid has previously been associated with increased systolic blood pressure (43). We also found a negative association between VAT fPUFA and T2D. VAT is known for its metabolic activity and association with features of the metabolic syndrome, such as hyperinsulinemia (44). Previous studies have shown that adipose tissue PUFA levels in mice reduce obesity-related insulin resistance (45). Further research involving longitudinal data could explore the FA composition tracking dietary and lifestyle changes, their impact on disease outcomes and their effects on adipose tissue.

While the study offers valuable insights, it is crucial to recognise its constraints. The UK Biobank, a substantial cross-sectional study, may be influenced by selection bias as it represents a ‘healthier’ group compared to the broader UK population and excludes younger individuals and cases of severe disease. Moreover, as the UK Biobank population is predominantly White, future work is needed to investigate diverse ethnic population data. Another potential limitation of this study is that the dietary parameters and inflammatory markers used were reported approximately 6.8 ± 1.5 years and 9 ± 1.7 years before the MRI acquisition, respectively. Finally, the relatively low R^2^ values suggest that many other factors contribute to FA composition; due to the observational nature of this analysis, relevant measurements such as adipocyte size, for instance, were not possible.

## Conclusion

Our large-scale MRI-based analysis of the FA composition of ASAT and VAT shows valuable insights into their associations with diet, anthropometrics, lifestyle, inflammation factors and disease. Our findings underscore the significance of assessing adipose tissue composition in relation to lifestyle and disease traits, paving the way for future research focusing on longitudinal and intervention studies to elucidate the impact of genetics and the exposome on adipose tissue composition and disease outcomes.

## FUNDING

This research was funded by a Calico Life Sciences LLC research grant.

## DISCLOSURE

None of the authors has a conflict of interest to disclose.

## AUTHOR CONTRIBUTIONS

J.D.B., E.L.T., and M.T. conceived the study. J.D.B., B.W., E.L.T., and M.T. designed the study. M.T., B.W. and N.B implemented the methods. M.T. performed the data and statistical analysis. E.L.T., B.W., M.T., J.D.B., and N.B. drafted the manuscript. All authors read and approved the manuscript.

## Acknowledgements

This research has been conducted using the UK Biobank Resource under Application Number 44584. We thank Calico Life Sciences LLC for funding our research.

## Declarations

Fully anonymised images and participant metadata were obtained through UK Biobank Access Application number 44584. The UK Biobank has approval from the North West Multi-Centre Research Ethics Committee (REC reference: 11/NW/0382), and obtained written informed consent from all participants before the study. All methods were performed in accordance with the relevant guidelines and regulations as presented by the appropriate authorities, including the Declaration of Helsinki.

## Data Availability

Our research was conducted using UK Biobank data. Under the standard UK Biobank data sharing agreement, we (and other researchers) cannot directly share raw data obtained or derived from the UK Biobank. However, under this agreement, all of the data generated, and methodologies used in this paper are returned by us to the UK Biobank, where they will be fully available. Access is obtained directly from the UK Biobank to all bona fide researchers upon submitting a health-related research proposal to the UK Biobank https://www.ukbiobank.ac.uk.

## Notes

### Competing Interest Statement

The authors have declared no competing interest.

