## Supplementary_Material for "MRI assessment of adipose tissue fatty acid composition in the UK Biobank and its association with diet and disease"

Disease Definitions

Categories of interest, including diet, hypertension, type-2 diabetes (T2D) and cardiovascular disease (CVD) were defined based on recorded hospital episode statistics (HES) data and self-reported information. If the International Classification of Diseases 10th edition for HES data, or self-reported disease codes (UK Biobank fields) were reported at least once for each participant, at the time before the first imaging visit, they were classified as a case. A summary of the codes corresponding to the considered disease traits are provided in supplementary Table S1. Hypertension was defined as self-report of hypertensive medication use, or a prior diagnosis of hypertension, or mean blood pressure ≥ 140/90 mmHg [3]. The codes for hypertension diagnosis were selected based on the ICD10 and the self-reported codes for hypertension. Hypertensive medication codes were selected from the medication fields, reporting as regularly taking blood pressure medication. The codes for T2D were selected based on the ICD10 and self-reported codes for type-2 diabetes (1). CVD was defined based on the ICD10 codes for angina, myocardial infarction, chronic ischaemic heart disease, atrial fibrillation, heart failure, and stroke (2).

Supplementary Tables

| **Trait** | **ICD-10** | **Self-Reported Code (20002)** | **UK Biobank Field** |
|---|---|---|---|

| | | | |
|---|---|---|---|
| **Diet** | - | - | 20086, 1389, 1369, 1359, 1349, 1379, 6144, 1418, 1408, 1428, 1329, 1339 |
| **Hypertension** | I10-I13, I15, O10 | 1065, 1072 | 6153, 6177 |
| **T2D** | E11 | 1220, 1223 | - |
| **CVD** | I20, I21, I25, I48, I50, I60, I61, I63, I64 | - | - |

Table S1. Summary of the codes used to define diet and disease.

| | **All (N = 33,583)** | **Women (N = 17,264)** | **Men (N = 16,319)** |
|---|---|---|---|
| | | **ASAT** | |
| **NDB** | 2.14 ± 0.24 | 2.19 ± 0.20 | 2.09 ± 0.27** |
| **NMIDB** | 0.46 ± 0.14 | 0.48 ± 0.11 | 0.45 ± 0.16** |
| | | **VAT** | |
| **NDB** | 2.12 ± 0.21 | 2.09 ± 0.23 | 2.16 ± 0.18** |
| **NMIDB** | 0.53 ± 0.11 | 0.52 ± 0.13 | 0.53 ± 0.10** |

Table S2. Summary statistics for NDB and NMIDB in ASAT and VAT of the full cohort (N = 33,583), separated by sex. Values are reported as mean and standard deviation. Significance refers to the p-value for a Wilcoxon rank-sums test, where the null hypothesis is the medians between the two groups are equal. ** indicate statistical significance between the vegan and omnivore participants, after Bonferroni correction (p = 0.00016). After Bonferroni correction (p = 0.00016), all comparisons between FA composition in ASAT and VAT were statistically significant.

Abbreviations: NDB, number of double bonds; NMIDB, number of methylene-interrupted double bonds; ASAT: abdominal subcutaneous adipose tissue; VAT: visceral adipose tissue.

| | **Vegan** | **Omnivore** |
|---|---|---|
| | **Women** | |
| **N** | 11 | 11 |
| **White (N)** | 10 | 11 |
| **Age (yrs.)** | 60.18 ± 6.82 | 73.09 ± 10.53* |
| **Weight (kg)** | 63.45 ± 10.52 | 97.40 ± 17.94* |
| **Height (cm)** | 163.04 ± 6.72 | 164.41 ± 6.39 |
| **BMI (kg/m²)** | 23.76 ± 3.14 | 35.93 ± 5.80* |
| **Walking MET (hours/week)** | 9.43 ± 9.87 | 7.25 ± 7.30 |
| **Moderate MET (hours/week)** | 18.09 ± 19.42 | 2.43 ± 3.93* |
| **Vigorous MET (hours/week)** | 5.20 ± 7.16 | 2.24 ± 6.31 |
| **Total MET (hours/week)** | 32.71 ± 26.35 | 11.92 ± 12.96* |
| **Townsend deprivation index** | 0.02 ± 2.41 | -2.99 ± 1.78* |
| **Alcohol frequency (N)** | | |

| | | |
|---|---|---|
| Daily | 2 | 2 |
| 1-4 times per week | 4 | 2 |
| 1-3 times per month | 1 | 2 |
| Occasional or never | 4 | 5 |
| **Smoking status (N)** | | |
| Never | 9 | 8 |
| Previous | 1 | 3 |
| Current | 1 | 0 |
| **Men** | | |
| **N** | 7 | 7 |
| **White (N)** | 7 | 7 |
| **Age (yrs.)** | 62.14 ± 8.05 | 69.14 ± 15.03* |
| **Weight (kg)** | 72.81 ± 8.23 | 106.40 ± 20.51* |
| **Height (cm)** | 176.36 ± 5.51 | 176.63 ± 6.40 |
| **BMI (kg/m²)** | 23.37 ± 1.96 | 34.09 ± 6.05* |
| **Walking MET (hours/week)** | 7.15 ± 5.96 | 10.10 ± 11.48 |
| **Moderate MET (hours/week)** | 31.39 ± 47.02 | 12.14 ± 17.25* |
| **Vigorous MET (hours/week)** | 12.75 ± 19.56 | 5.25 ± 11.65 |
| **Total MET (hours/week)** | 51.29 ± 68.69 | 27.50 ± 34.62* |
| **Townsend deprivation index** | 0.21 ± 3.51 | -1.66 ± 2.67* |

| | | |
|---|---|---|
| **Alcohol frequency (N)** | | |
| **Daily** | 1 | 1 |
| **1-4 times /week** | 2 | 3 |
| **1-3 times/month** | 1 | 1 |
| **Occasional or never** | 3 | 2 |
| **Smoking status (N)** | | |
| **Never** | 4 | 3 |
| **Previous** | 3 | 3 |
| **Current** | 0 | 1 |

Table S3. Demographics of the vegan and omnivore participants, separated by sex. Values are reported as mean and standard deviation for continuous variables and counts (N) for categorical variables. Significance refers to the p-value for a Wilcoxon rank-sums test, where the null hypothesis is the medians between the two groups (vegan and omnivore participants) being equal. An asterisk (*) indicates statistically significant for p-value < 0.05.

Abbreviations: BMI, body mass index; MET, metabolic equivalents of task.

| | **Vegan (N = 18)** | **Omnivore (N = 18)** |
|---|---|---|
| | **ASAT** | |
| **NDB** | 2.66 ± 0.17 | 2.35 ± 0.58** |
| **NMIDB** | 0.69 ± 0.10 | 0.59 ± 0.39* |
| | **VAT** | |

| | | |
|---|---|---|
| **NDB** | 2.74 ± 0.32 | 2.27 ± 0.18** |
| **NMIDB** | 0.81 ± 0.21† | 0.55 ± 0.09** |

Table S4. Summary statistics for NDB and NMIDB in ASAT and VAT of the vegan and omnivore participants (N = 36). Values are reported as mean and standard deviation. Significance refers to the p-value for a Wilcoxon rank-sums test, where the null hypothesis is the medians between the two groups are equal. An asterisk (*) indicates statistically significant for p-value < 0.05, ** indicate statistical significance between the vegan and omnivore participants, after Bonferroni correction (p = 0.00016). † indicates statistical significance between FA composition in ASAT and VAT, for p-value < 0.05.

Abbreviations: NDB, number of double bonds; NMIDB, number of methylene-interrupted double bonds; ASAT: abdominal subcutaneous adipose tissue; VAT: visceral adipose tissue.

| | ASAT fSFA | ASAT fMUFA | ASAT fPUFA | VAT fSFA | VAT fMUFA | VAT fPUFA |
|---|---|---|---|---|---|---|
| Age | -0.096 | -0.315 | 0.264 | 0.086 | -0.158 | 0.018 |
| | (0.180) | (0.197) | (0.221) | (0.159) | (0.240) | (0.199) |
| Sex [Men] | 0.135 | -0.069 | -0.038 | -0.423 | 0.091 | 0.251 |
| | (0.297) | (0.325) | (0.364) | (0.263) | (0.395) | (0.327) |
| Alcohol frequency [Daily] | 0.122 | 0.016 | -0.081 | 0.276 | 0.160 | -0.276 |
| | (0.426) | (0.467) | (0.523) | (0.377) | (0.567) | (0.470) |
| Alcohol frequency [1-4 times/week] | -0.583 | -0.807* | 0.903* | 0.237 | -0.534 | 0.115 |
| | (0.340) | (0.373) | (0.418) | (0.301) | (0.453) | (0.375) |
| Alcohol frequency [1-3 times/month] | -0.518 | -0.314 | 0.531 | -0.179 | 0.240 | -0.006 |
| | (0.483) | (0.529) | (0.593) | (0.428) | (0.643) | (0.533) |
| Smoking status [Current] | 0.147 | -0.188 | 0.030 | 0.902 | -0.579 | -0.346 |
| | (0.643) | (0.705) | (0.790) | (0.570) | (0.856) | (0.710) |
| Smoking status [Previous] | 0.040 | -0.429 | 0.272 | 0.297 | -0.425 | 0.012 |
| | (0.339) | (0.372) | (0.417) | (0.300) | (0.451) | (0.374) |
| MET | -0.015 | -0.185 | 0.137 | 0.036 | -0.033 | -0.008 |
| | (0.157) | (0.172) | (0.193) | (0.139) | (0.209) | (0.173) |
| Townsend | -0.229 | -0.27 | 0.323 | 0.250 | -0.078 | -0.136 |

| | | | | | | |
|---|---|---|---|---|---|---|
| **deprivation index** | (0.162) | (0.178) | (0.199) | (0.144) | (0.216) | (0.179) |
| **Omnivore** | 1.194* | -0.714 | -0.223 | **1.556**** | 0.596 | -1.410* |
| | (0.359) | (0.393) | (0.441) | **(0.318)** | (0.478) | (0.396) |
| **Constant** | -0.439 | 0.801* | -0.287 | -0.840* | -0.08 | 0.635 |
| | (0.284) | (0.311) | (0.348) | (0.251) | (0.377) | (0.313) |
| **Observations** | 36 | 36 | 36 | 36 | 36 | 36 |
| **Adjusted $R^2$** | 0.388 | 0.266 | 0.077 | 0.520 | -0.084 | 0.255 |
| **AIC** | 95.4 | 101.9 | 110.2 | 86.6 | 115.9 | 102.4 |

Table S5. Summary of standardised regression coefficients (β) for linear regression models of fatty acid composition in ASAT and VAT representing the associations with baseline characteristics on the vegan and omnivore participants (N = 36). Standard errors are shown in parentheses. An asterisk (*) indicates statistically significant for p-value < 0.05, ** indicate statistically significant after Bonferroni correction (p = 0.00016). Significant associations after Bonferroni correction are shown in bold.

Abbreviations: fSFA, saturated fatty acid fraction; fPUFA, polyunsaturated fatty acids fraction; fMUFA, monounsaturated fatty acids fraction; MET, metabolic equivalents of task; ASAT: abdominal subcutaneous adipose tissue; VAT: visceral adipose tissue; AIC, Akaike information criterion.

| | **ASAT fSFA** | **ASAT fMUFA** | **ASAT fPUFA** | **VAT fSFA** | **VAT fMUFA** | **VAT fPUFA** |
|---|---|---|---|---|---|---|
| **Age** | **0.048**** | -0.020* | -0.027* | 0.005 | **0.034**** | -0.036* |

| | | | | | | |
|---|---|---|---|---|---|---|
| | **(0.009)** | (0.009) | (0.010) | (0.008) | **(0.008)** | (0.010) |
| **Sex [Men]** | **0.468**** | **-0.264**** | **-0.209**** | **-0.492**** | **0.480**** | **0.110**** |
| | **(0.018)** | **(0.019)** | **(0.019)** | **(0.017)** | **(0.017)** | **(0.019)** |
| **Ethnicity [Asian]** | **-0.557**** | **0.419**** | 0.161 | **-0.716**** | **0.342**** | **0.462**** |
| | **(0.080)** | **(0.083)** | (0.084) | **(0.072)** | **(0.073)** | **(0.084)** |
| **Ethnicity [Black]** | -0.385* | -0.125 | 0.411* | -0.027 | **-0.552**** | **0.515**** |
| | (0.125) | (0.129) | (0.132) | (0.113) | **(0.114)** | **(0.132)** |
| **Ethnicity [Chinese]** | **-0.741**** | 0.564* | 0.211 | -0.470* | -0.089 | 0.578* |
| | **(0.164)** | (0.169) | (0.173) | (0.148) | (0.149) | (0.172) |
| **Ethnicity [Others]** | -0.319* | 0.151 | 0.155 | -0.244* | 0.063 | 0.200* |
| | (0.092) | (0.096) | (0.098) | (0.084) | (0.084) | (0.097) |
| **BMI** | **-0.137**** | **0.200**** | -0.021* | **-0.399**** | **0.385**** | **0.094**** |
| | **(0.009)** | **(0.010)** | (0.010) | **(0.008)** | **(0.009)** | **(0.010)** |
| **Alcohol frequency [Daily]** | **0.200**** | -0.020 | **-0.149**** | 0.080* | 0.035 | -0.112* |
| | **(0.030)** | (0.031) | **(0.032)** | (0.027) | (0.027) | (0.032) |
| **Alcohol frequency [1-4 times/week]** | **0.133**** | 0.014 | **-0.119**** | 0.027 | -0.005 | -0.020 |
| | **(0.024)** | (0.025) | **(0.026)** | (0.022) | (0.022) | (0.025) |
| **Alcohol frequency [1-3 times/month]** | 0.038 | 0.059 | -0.071* | -0.004 | 0.009 | 0.0003 |
| | (0.032) | (0.033) | (0.034) | (0.029) | (0.029) | (0.034) |

| | | | | | | |
|---|---|---|---|---|---|---|
| **Smoking status** | -0.015 | -0.115* | 0.090 | -0.011 | -0.016 | 0.026 |
| **[Current]** | (0.052) | (0.054) | (0.055) | (0.047) | (0.048) | (0.055) |
| **Smoking status** | -0.001 | -0.010 | 0.007 | -0.002 | 0.019 | -0.013 |
| **[Previous]** | (0.019) | (0.019) | (0.020) | (0.017) | (0.017) | (0.020) |
| | **0.035**** | **-0.052**** | 0.007 | **0.068**** | **-0.072**** | -0.010 |
| **Vigorous MET** | **(0.009)** | **(0.009)** | (0.009) | **(0.008)** | **(0.008)** | (0.009) |
| **Townsend** | -0.026* | -0.013 | 0.029* | -0.002 | 0.020* | -0.016 |
| **deprivation index** | (0.009) | (0.009) | (0.009) | (0.008) | (0.008) | (0.009) |
| | **-0.210**** | **0.101**** | **0.102**** | **-0.190**** | **0.101**** | **0.113**** |
| **Dietary MUFA** | **(0.015)** | **(0.015)** | **(0.016)** | **(0.013)** | **(0.014)** | **(0.016)** |
| | **0.303**** | **-0.129**** | **-0.160**** | **0.255**** | **-0.090**** | **-0.191**** |
| **Dietary SFA** | **(0.014)** | **(0.015)** | **(0.015)** | **(0.013)** | **(0.013)** | **(0.015)** |
| | **-0.049**** | -0.006 | **0.044**** | -0.029* | -0.033* | **0.059**** |
| **Carbohydrate** | **(0.011)** | (0.011) | **(0.011)** | (0.010) | (0.010) | **(0.011)** |
| | 0.016 | -0.014 | -0.004 | 0.014 | -0.016 | -0.002 |
| **Protein** | (0.011) | (0.012) | (0.012) | (0.010) | (0.010) | (0.012) |
| | 0.020* | -0.003 | -0.014 | 0.002 | 0.029* | -0.029* |
| **CRP** | (0.009) | (0.009) | (0.009) | (0.008) | (0.008) | (0.009) |
| **WBCC** | 0.004 | 0.018* | -0.015 | **-0.039**** | **0.045**** | 0.003 |

| | | | | | | |
|---|---|---|---|---|---|---|
| | (0.009) | (0.009) | (0.009) | **(0.008)** | **(0.008)** | (0.009) |
| **Hypertension** | 0.001 | -0.038 | 0.026 | -0.058* | **0.076**** | -0.004 |
| | (0.019) | (0.020) | (0.020) | (0.017) | **(0.018)** | (0.020) |
| **T2D** | 0.121* | -0.110* | -0.023 | 0.110* | 0.081* | **-0.191**** |
| | (0.039) | (0.041) | (0.042) | (0.036) | (0.036) | **(0.042)** |
| **CVD** | -0.061* | 0.033 | 0.028 | -0.026 | 0.027 | 0.0004 |
| | (0.030) | (0.031) | (0.031) | (0.027) | (0.027) | (0.031) |
| **Constant** | **-0.327**** | **0.134**** | **0.178**** | **0.243**** | **-0.282**** | -0.020 |
| | **(0.023)** | **(0.024)** | **(0.024)** | **(0.021)** | **(0.021)** | (0.024) |
| **Observations** | 12,128 | 12,128 | 12,128 | 12,128 | 12,128 | 12,128 |
| **Adjusted $R^2$** | 0.131 | 0.069 | 0.029 | 0.289 | 0.277 | 0.035 |
| **AIC** | 32744 | 33573.9 | 34085.2 | 30305.1 | 30504.8 | 34017.1 |

Table S6. Summary of regression coefficients (β) for linear regression models of the fatty acid composition in ASAT and VAT representing the associations with baseline characteristics on the full cohort (N=13,176). Standard errors are shown in parentheses. An asterisk (*) indicates statistically significant for p-value < 0.05, ** indicate statistically significant after Bonferroni correction (p = 0.00016). Significant associations after Bonferroni correction are shown in bold.

Abbreviations: fSFA, saturated fatty acid fraction; fPUFA, polyunsaturated fatty acids fraction; fMUFA, monounsaturated fatty acids fraction; BMI, body mass index; MET, metabolic equivalents of task; FA, fatty acid; CRP, c-reactive protein; WBCC, white blood cell count; T2D, type-2 diabetes; CVD, cardiovascular disease; ASAT: abdominal subcutaneous adipose tissue; VAT: visceral adipose tissue; AIC, Akaike information criterion.

Supplementary Figures

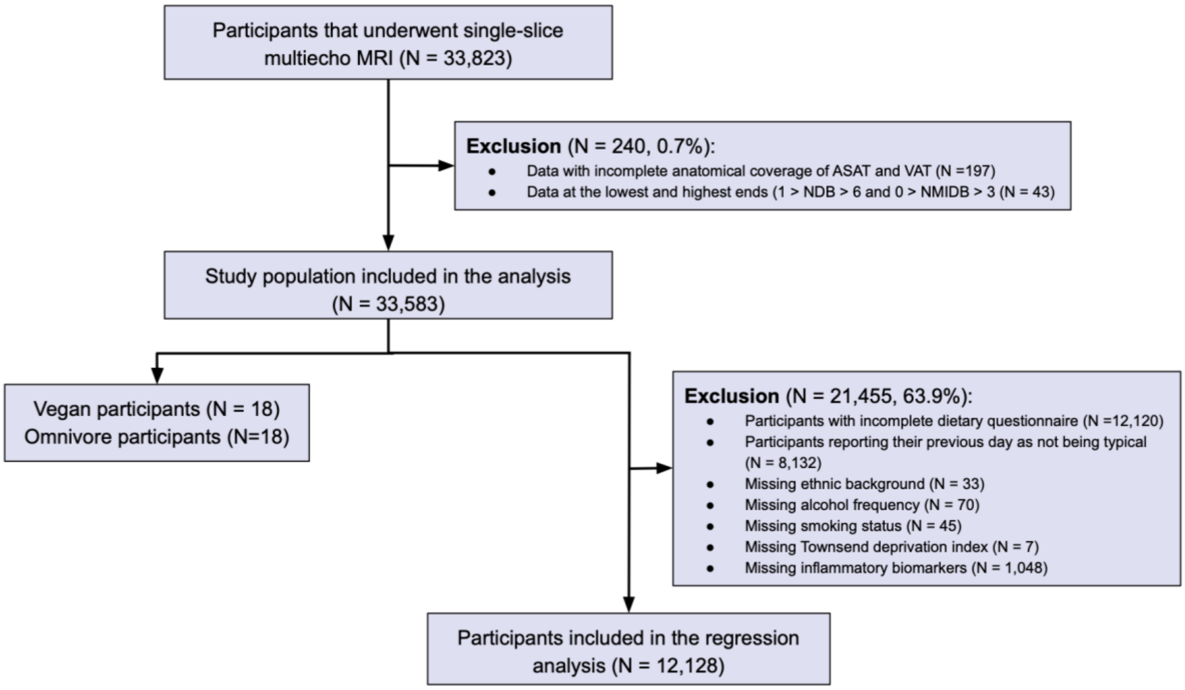


Figure S1. Flow diagram of the study population.
